# Practice Variation in Vasoactive Medication Use for Cardiogenic Shock Across ICU Subtypes: A Retrospective Cohort Study

**DOI:** 10.1101/2025.07.11.25331408

**Authors:** Jackson L Shriver, David E Hamilton, Ashley M Hesson, Michael R Mathis, Andrea D Thompson

**Affiliations:** University of Michigan, Department of Internal Medicine, Ann Arbor, MI; University of Michigan, Department of Internal Medicine, Division of Cardiovascular Medicine, Ann Arbor, MI; University of Michigan, Department of Obstetrics & Gynecology, Division of Maternal Fetal Medicine, Ann Arbor, MI; University of Michigan, Department of Anesthesia, Ann Arbor, MI

## Abstract

**BACKGROUND:** Cardiogenic shock has significant associated morbidity and mortality with a wide range of vasoactive management strategies. However, the extent to which variation in vasoactive and inodilator therapies is explained by patient level variables versus clinical practice variation remains underexplored.

**METHODS:** The cohort included 4,525 patients admitted to ICUs at Michigan Medicine from 01/01/2014–12/31/2023 diagnosed with cardiogenic shock identified by billing codes. Vasoactive medication utilization was compared across various ICUs (cardiovascular, cardiovascular surgical, non-cardiac). Mixed-effect multiple logistic modeling was used to evaluate to what extent variation in inodilator use was associated with fixed patient-level variables (i.e. prior cardiac arrest) compared to ICU location as a random effect.

**RESULTS:** Patient encounters were classified as cardiovascular ICU (n = 1,355), cardiovascular surgical ICU (n = 1,405), non-cardiac ICU (n = 723), and multiple ICUs (n = 1,042). Vasoactive and inodilator medication use varied significantly. Inodilators were more frequently used in the cardiac ICU (44.4% [95% CI 40.4-48.3%]), cardiothoracic surgical ICU (64.9% [61.8-68.0%]), and multiple ICU patients (60.8% [57.0-64.6%]) and less frequently in non-cardiac ICU patients (18.8% [12.2-25.4%]). Heart failure was associated with more frequent inodilator use (OR 3.82 [3.06-4.80]), while increased age (OR 0.993 [0.989 – 0.997]), male sex (0.80 [0.70 – 0.91]), and prior cardiac arrest (0.68 [0.58 – 0.81]) were associated with lower use. ICU location attributed to 15.8% of variance in inodilator use, while fixed patient-level variables combined accounted for 6.1% of variance.

**CONCLUSION:** A substantial portion of variation in vasoactive medication utilization was attributed to ICU setting.

## Introduction

Despite significant advances in the treatment of cardiogenic shock, it remains a highly morbid condition with mortality estimated as high as 30-40%.^1–3^ The mainstays of treatment involve the use of a combination of vasoactive medications and mechanical circulatory support (MCS) to manage the complex pathophysiology of cardiogenic shock, as well as targeted treatment to address the underlying etiology of shock. These vasoactive medication treatment options range from vasopressors, vasodilators, inodilators, and inopressors with different impacts on vascular tone and cardiac contractility. However, clinical trials comparing vasoactive therapies for the treatment of cardiogenic shock are limited.^4–7^ Professional society scientific statements and guidelines highlight this lack of high-quality data, thereby avoiding firm recommendations for first-line vasoactive agents and inotropes.^2,8–11^ We hypothesize that significant variation exists in the use of vasoactive medications for treating cardiogenic shock across different types of intensive care units (ICUs).

This hypothesis is supported by prior work that has highlighted variability between institutions for multiple cardiogenic shock interventions, including the use of pulmonary artery catheters and mechanical circulatory support devices (MCS).^3,12^ Further, studies in cardiac surgery populations have shown that variability in vasoactive medication utilization is influenced by provider-level, institution-level, and patient-level factors.^13^ Variability in vasoactive medication use could be driven by precision medicine, tailoring therapy to specific pathophysiology. Alternatively, variability could be driven by clinical equipoise or differences in culture, resource availability, provider training, or experience. However, an understanding of the variation in vasoactive medication use in the treatment of cardiogenic shock remains lacking. Within a single institution, different ICUs, staffed by critical care providers with different medical training (e.g., pulmonary, surgery, anesthesia, cardiology), may care for patients with cardiogenic shock differently. To date, this practice pattern variation has not been defined, and it is unclear what drives underlying variability, and whether variation leads to improvement in patient care, or if further standardization may lead to improved outcomes.

Thus, we aim to characterize current vasoactive utilization patterns across various ICUs at a single institution. We will define the factors associated with variability in vasoactive medication utilization as a first step in developing strategies to study differences in the effectiveness of vasoactive medication use in treating cardiogenic shock. Understanding the drivers of variation can lead to future efforts to optimize clinical care and allow for practice standardization when appropriate.

## Methods

### Study Design and Patient Selection

This is a retrospective cohort study of adult patients admitted to intensive care units (ICUs) at Michigan Medicine between January 1, 2014 and December 31, 2023. Eligible patients were aged between 18 to 100 years and diagnosed with cardiogenic shock, defined by use of the ICD 9 code 785.51 or ICD 10 code R57.0.^14–18^ Patients were identified using Data Direct, a clinical data repository that allowed for collection of admission date, unit of admission, fixed patient-level variables such as; demographics, comorbidities, laboratory results, medication utilization, left ventricular ejection fraction (LVEF), use of mechanical circulatory support (MCS), and hospital outcomes.

Patients were subclassified into specific ICU subgroups based on patient locations identified during their hospital encounter. Patients were classified as being in cardiac ICU, cardiothoracic surgical ICU, and non-cardiac ICUs. The non-cardiac ICU group was composed of a combination of four separate ICU environments, including the surgical ICU, two different medical ICUs, and the neurocritical care unit. Patients who were cared for in multiple of these groups during one hospital admission were reclassified into a fourth group referred to as a multiple ICU cohort.

### Data Collection

For patients with cardiogenic shock, medication administration during the patient’s ICU stay was reviewed for eight identified vasoactive medications: norepinephrine, epinephrine, phenylephrine, dopamine, vasopressin, dobutamine, milrinone, and nitroprusside. Single-dose and non-infusion doses were excluded. Inodilator use was defined as any use of either milrinone or dobutamine during the patient’s ICU stay.

Comorbidities for each patient were identified using the Elixhauser Comorbidity Index,^19,20^ supplemented by additional ICD 9/10 codes for dialysis, cardiac arrest, septic shock, hypovolemic shock, and ventricular tachycardia. MCS was identified by using procedure codes for the use of intra-aortic balloon pumps, extracorporeal membrane oxygenation (ECMO), percutaneous external heart assist systems, percutaneous ventricular assistance devices, and temporary microaxial devices to augment cardiac output. The mean LVEF was calculated by averaging the LVEFs obtained during the hospital admission.

The protocol was reviewed by the University of Michigan Institutional Review Board and was approved (HUM00252222).

### Statistical Methods

Non-parametric descriptive statistics and testing methods were primarily used based on the data distributions across our variables of interest. Continuous variables were evaluated with medians and interquartile ranges (IQR). Categorical variables were compared as counts. Comparison between groups of continuous variables was performed using the Wilcoxon rank-sum test and Kruskal-Wallis tests. Categorical variables were compared using Chi-Square tests. Dot density plots were generated by plotting one point per 0.1% utilization across each ICU subtype.

Mixed-effect multiple logistic modeling was used to evaluate variables driving inodilator utilization. Fixed effects in the model included patient-level variables of age, sex, preceding cardiac arrest, septic shock, hypovolemic shock, cardiac arrhythmia, and congestive heart failure. Resultant states (ie hypovolemic shock, septic shock, and congestive heart failure) were found to have covariation with predisposing factors (acute blood loss anemia, positive blood cultures, and low LVEF); thus, predisposing factors were removed from analysis. Mechanical circulatory support utilization was initially compared but excluded from analysis, as usage was unequally distributed across ICU environments. ICU type was included as the random effect, and inodilator use was the dependent variable. Analysis was performed using R 4.41 (R Foundation for Statistical Computing, Vienna, Austria). Statistical significance was assessed at a p-value of 0.05.

## Results

### ICU Demographics

A total of 5,717 ICU admissions were identified across 3,977 unique patients diagnosed with cardiogenic shock. After excluding patients with no ICU encounter (n = 31) and hospital admissions outside of the study period (n = 39), a total of 4,525 hospital encounters across 3,946 patients were included (Figure 1).

**Figure 1:**
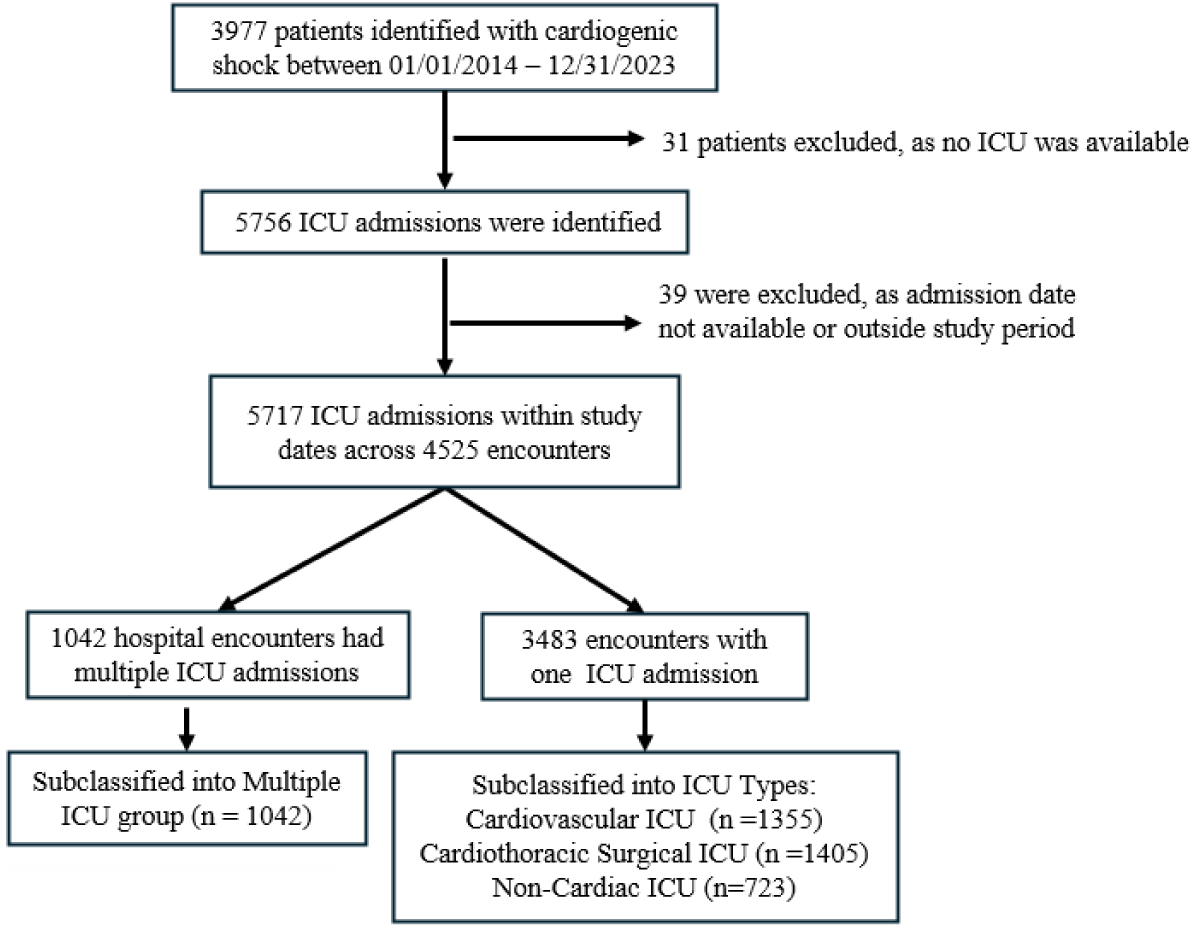
Cohort identification and patient exclusion criteria.

Patients were subsequently classified into ICU subgroups: cardiac ICU (n = 1,355), cardiothoracic surgical ICU (n = 1,405), non-cardiac ICU (n = 723), and multiple ICU cohort (n = 1,042). The median age of the identified patients was 62 years (IQR, 51 –71). Overall, 65.5% of patients were male. The population was predominantly Caucasian (75.6%), with 17.3% African American, 1.7% Asian American, and smaller percentages of other racial groups.

Several notable demographic differences are observed across ICUs. For example, a younger median age was identified in patients within the multiple ICU group (59 years, IQR 47 – 68) and cardiothoracic surgical ICU (62 years, IQR 51-71), and older patients were identified in the cardiac ICU (63 years IQR 51-71) and non-cardiac ICU (64 years, IQR 53-74). Median LVEF was 37.5% and was lowest in cardiac ICU patients (26.3%, IQR 18-48) and multiple ICU patients (36.4%, IQR 18%–60 %). LVEF values were missing in 1,549 patients, including all patients who were admitted before May of 2015. Mechanical circulatory support (MCS) was used in 23.2% of patients, most commonly in multiple ICU patients (40.4%) and cardiothoracic surgical ICU patients (28.6%). Concomitant septic shock was more common in non-cardiac ICU populations (53.3%) and multiple ICU patients (30.3%) (Table 1).

**Table 1:** Demographic Table and Selected Comorbidities

|  | **Overall** | **Cardiac ICU** | **Cardiothoracic**<br>**Surgical ICU** | **Non-Cardiac ICU** | **Multiple ICU** | **p** |
| --- | --- | --- | --- | --- | --- | --- |
| n | 4525 | 1355 | 1405 | 723 | 1042 |  |
| Age (median [IQR]) | 62 [51, 71] | 63.00 [53, 73] | 62.00 [53, 71] | 64.00 [53, 74] | 59.00 [47.3, 68] | <0.001 |
| Sex, Male (%) | 2966 (65.5) | 917 (67.7) | 927 (66.0) | 416 (57.5) | 706 (67.8) | <0.001 |
| Race (%) |  |  |  |  |  | 0.004 |
| African American | 774 (17.3) | 244 (18.3) | 211 (15.2) | 103 (14.4) | 216 (20.9) |  |
| American Indian | 24 (0.5) | 6 (0.5) | 11 (0.8) | 3 (0.4) | 4 (0.4) |  |
| Asian | 77 (1.7) | 22 (1.7) | 21 (1.5) | 23 (3.2) | 11 (1.1) |  |
| Caucasian | 3378 (75.6) | 1007 (75.6) | 1069 (77.1) | 550 (77.1) | 752 (72.7) |  |
| Native Hawaiian and Other Pacific<br>Islander | 4 (0.1) | 1 (0.1) | 3 (0.2) | 0 (0.0) | 0 (0.0) |  |
| Other | 116 (2.6) | 24 (1.8) | 39 (2.8) | 20 (2.8) | 33 (3.2) |  |
| Patient Refused | 11 (0.2) | 4 (0.3) | 4 (0.3) | 1 (0.1) | 2 (0.2) |  |
| Unknown | 83 (1.9) | 24 (1.8) | 29 (2.1) | 13 (1.8) | 17 (1.6) |  |
| BMI (median [IQR]) | 27.5 [23.6, 32.2] | 26.8 [23. 31] | 28.5 [24.8, 33.3] | 27.5 [23.3, 33.2] | 27.07 [23.13, 32.04] | <0.001 |
| Ejection Fraction (median [IQR]) | 37.5 [20.0, 60.0] | 26.3 [18.0, 48.4] | 50.0 [20.0, 62.5] | 55.0 [30.9, 65] | 36.4 [18.3, 60] | <0.001 |
| Ventricular Tachycardia (%) | 1245 (27.5) | 434 (32.0) | 331 (23.6) | 97 (13.4) | 383 (36.8) | <0.001 |
| Cardiac Arrest (%) | 808 (17.9) | 249 (18.4) | 203 (14.4) | 138 (19.1) | 218 (20.9) | <0.001 |
| Septic Shock (%) | 1148 (25.4) | 251 (18.5) | 196 (14.0) | 385 (53.3) | 316 (30.3) | <0.001 |
| Hypovolemic Shock (%) | 152 (3.4) | 21 (1.5) | 50 (3.6) | 35 (4.8) | 46 (4.4) | <0.001 |
| Dialysis (%) | 669 (14.8) | 154 (11.4) | 200 (14.2) | 137 (18.9) | 178 (17.1) | <0.001 |
| Alcohol Abuse (%) | 432 (9.5) | 136 (10.0) | 120 (8.5) | 85 (11.8) | 91 (8.7) | 0.075 |
| Acute Blood Loss Anemia (%) | 892 (19.7) | 177 (13.1) | 309 (22.0) | 186 (25.7) | 220 (21.1) | <0.001 |
| Cardiac Arrhythmia (%) | 4173 (92.2) | 1264 (93.3) | 1269 (90.3) | 651 (90.0) | 989 (94.9) | <0.001 |
| Chronic Pulmonary Disease (%) | 2442 (54.0) | 728 (53.7) | 759 (54.0) | 382 (52.8) | 573 (55.0) | 0.839 |
| Congestive Heart Failure (%) | 3950 (87.3) | 1274 (94.0) | 1200 (85.4) | 525 (72.6) | 951 (91.3) | <0.001 |
| Diabetes (%) | 2049 (45.3) | 680 (50.2) | 582 (41.4) | 325 (45.0) | 462 (44.3) | <0.001 |
| Hypertension (%) | 3754 (83.0) | 1152 (85.0) | 1182 (84.1) | 587 (81.2) | 833 (79.9) | 0.003 |
| Liver Disease (%) | 2003 (44.3) | 610 (45.0) | 544 (38.7) | 366 (50.6) | 483 (46.4) | <0.001 |
| Metastatic Cancer (%) | 658 (14.5) | 193 (14.2) | 154 (11.0) | 172 (23.8) | 139 (13.3) | <0.001 |
| Obesity (%) | 1813 (40.1) | 531 (39.2) | 593 (42.2) | 274 (37.9) | 415 (39.8) | 0.207 |
| Peripheral Vascular Disease (%) | 2856 (63.1) | 851 (62.8) | 988 (70.3) | 337 (46.6) | 680 (65.3) | <0.001 |
| Valvular Disease (%) | 2888 (63.8) | 865 (63.8) | 1075 (76.5) | 268 (37.1) | 680 (65.3) | <0.001 |
| Mechanical Circulatory Support (%) | 1049 (23.2) | 189 (13.9) | 402 (28.6) | 35 (4.8) | 423 (40.6) | <0.001 |

### Vasoactive Medication Utilization

In 91.5% of patients, at least one vasoactive medication was utilized during their admission. The median number of distinct vasoactive medications per patient was found to be three (IQR 2-4). The total number of vasoactive medications used varied significantly among ICU types (p-value < 0.001), with a higher number of vasoactive medications utilized among the cardiothoracic surgical and the multiple ICU group (Figure 2).

**Figure 2:**
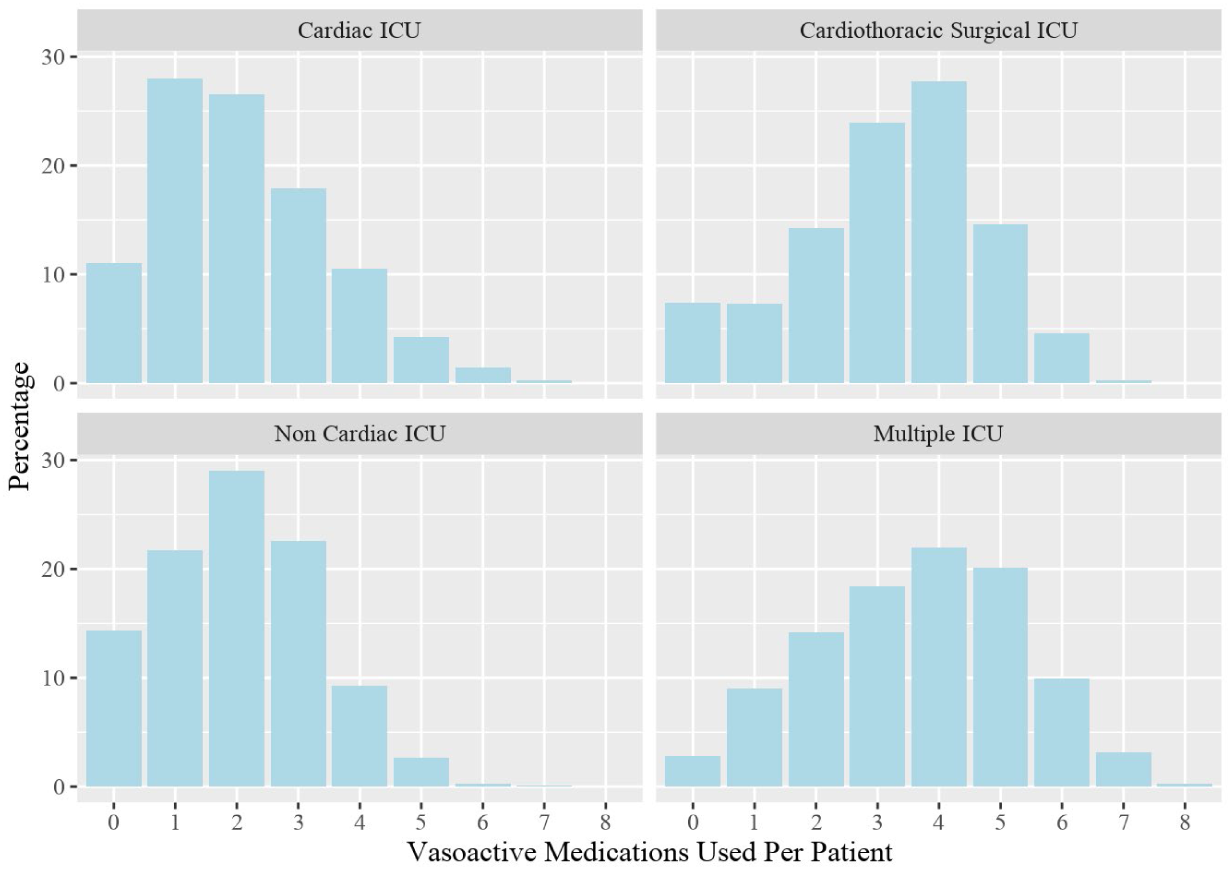
Number of vasoactive medication infusions per patent by ICU type

There was also significant variation in the frequency of use for individual vasoactive medications across ICUs (Figure 3). Utilization of each vasoactive medication varied significantly (p-value <0.001) across ICU subtypes. Norepinephrine was the most used vasoactive medication (77.1%) with variability across ICUs ranging from the cardiac ICU (61.4%) to the multiple ICU cohort (85.8%). Epinephrine was used in 42.1% of encounters, most commonly in the cardiothoracic surgical ICU (61.4%) and least commonly in the cardiac ICU (13.8%).

**Figure 3:**
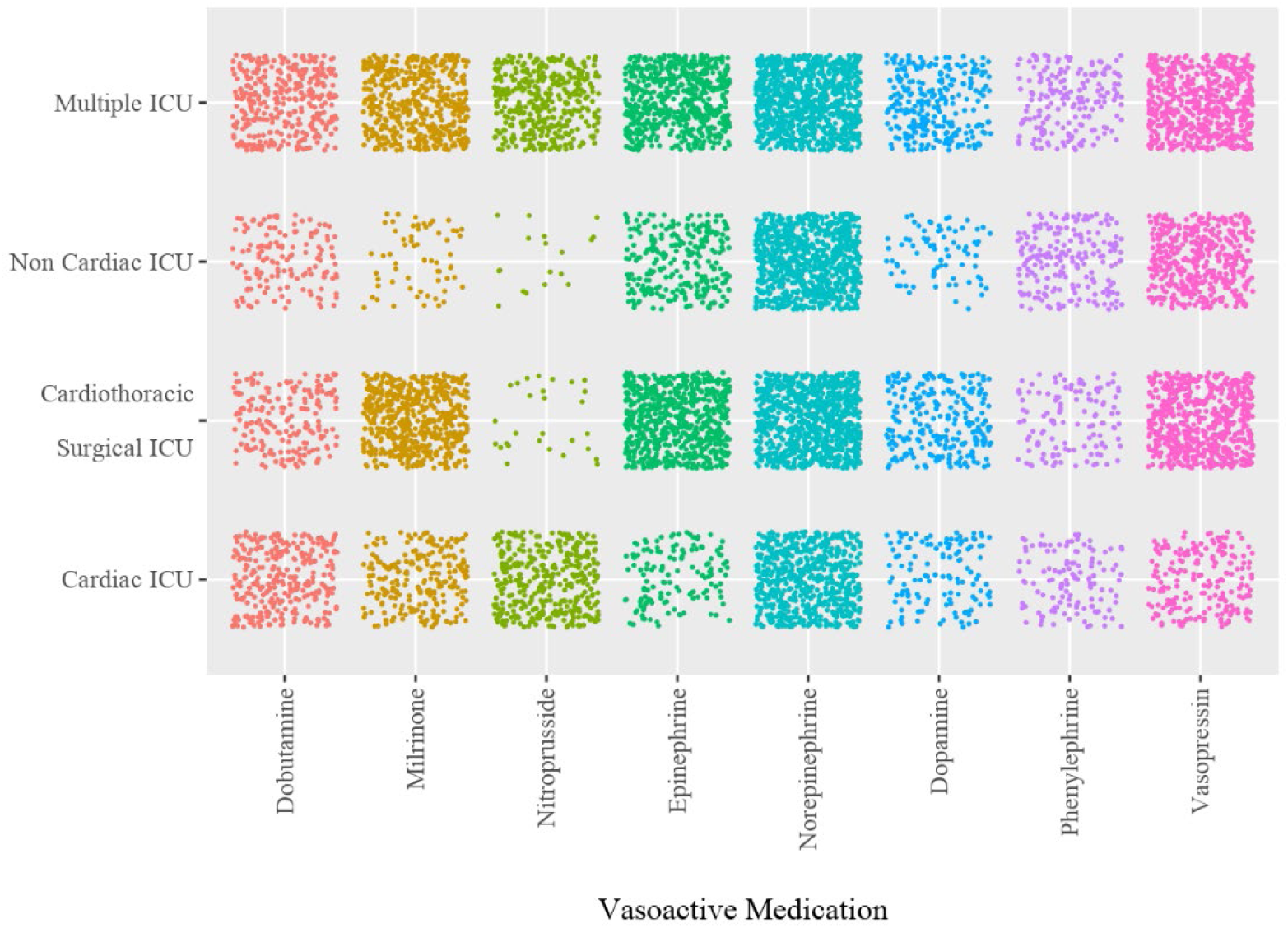
Dot density plot demonstrating vasoactive medication utilization rate. Each plotted point equals 0.1% utilization rate of each vasoactive medication.

### Inodilator Utilization

The use of individual inodilators, either dobutamine or milrinone, varied significantly (p-value < 0.001) across ICUs. Dobutamine was used in 24.8% of patient encounters and was most commonly used in multiple ICU (34.0%) patients and cardiac ICU (29.2%) patients. Dobutamine was less frequently used in non-cardiac ICU (13.8%) and cardiothoracic surgical ICU (19.4%) patients. Milrinone was utilized more regularly, at a rate of 35.8% overall, with the highest use in the cardiothoracic surgical ICU (55.1%) and multiple ICU (47.3%) groups, and less frequently in cardiac ICU (22.7%) and non-cardiac ICU (6.2%) patients.

This variation persisted when analyzing the use of inodilators as a whole. At least one inodilator was used in 50.5% of patients, and the use of inodilators was significantly different across ICUs (p-value < 0.001). Use was most frequent in cardiothoracic surgical ICU patients (64.9%), followed by multiple ICU patients (60.8%), and was used less commonly in cardiac ICU patients (44.4%) and dramatically less in non-cardiac ICU patients (18.8%) (Figure 4).

**Figure 4:**
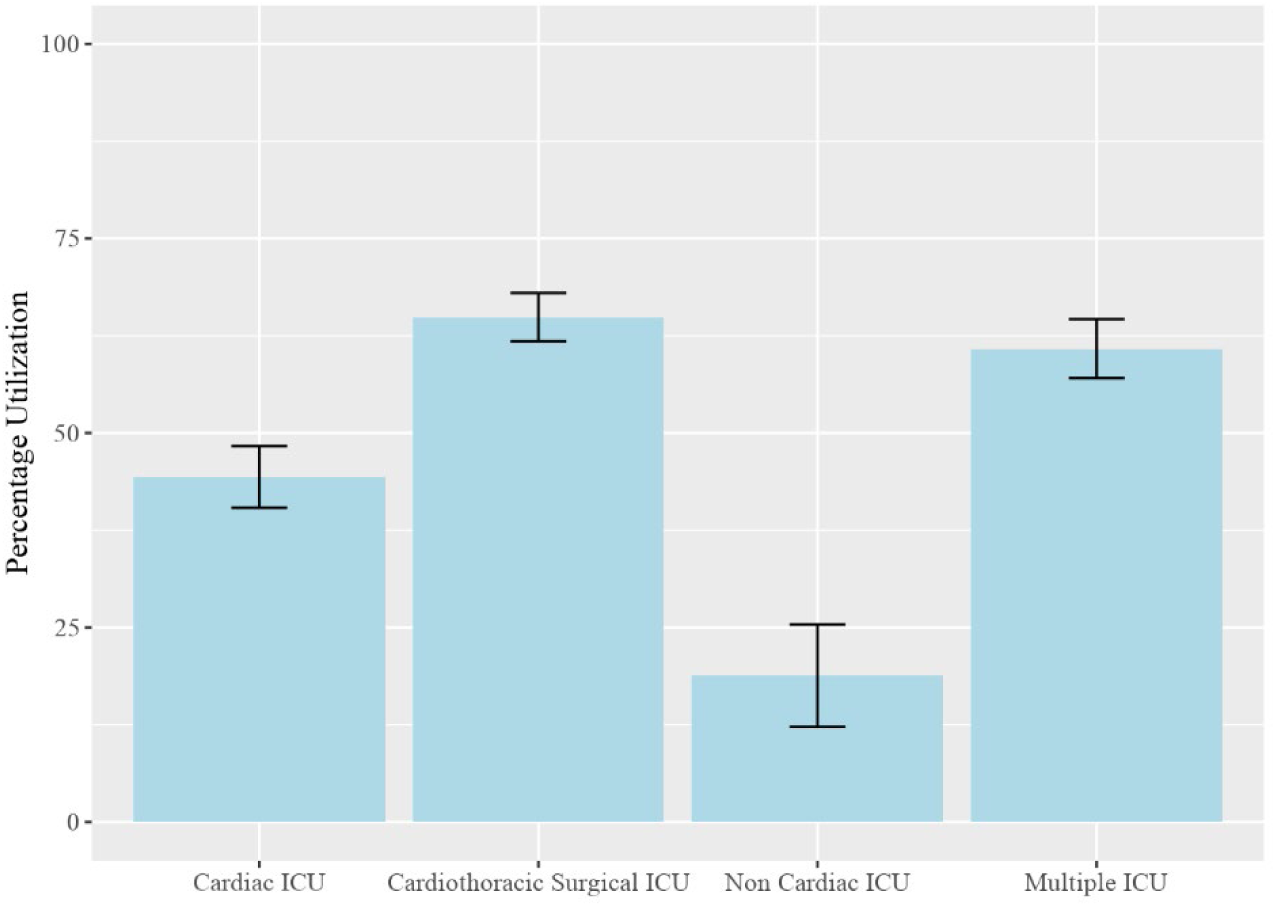
Proportion of patients who received at least one inodilator medication

### Identifying Factors Associated with Inodilator Use with Mixed Effect Modeling

Mixed effect modeling (Figure 5) revealed that a history of congestive heart failure was most strongly associated with increased use of inodilators (OR [95% CI]: 3.82 [3.06 – 4.80]). Less frequent inodilator use was associated with male sex (0.80 [0.70 – 0.91]) and cardiac arrest (0.68 [0.58 – 0.81]). Increasing age was mildly associated with less inodilator use (0.993 [0.989-0.997]). Concomitant septic shock (0.88 [0.75 – 1.02]) and hypovolemic shock (0.88 [0.61-1.26]), and cardiac arrhythmia (1.05 [0.82-1.34]) were not associated with inodilator use (Figure 5).

**Figure 5:**
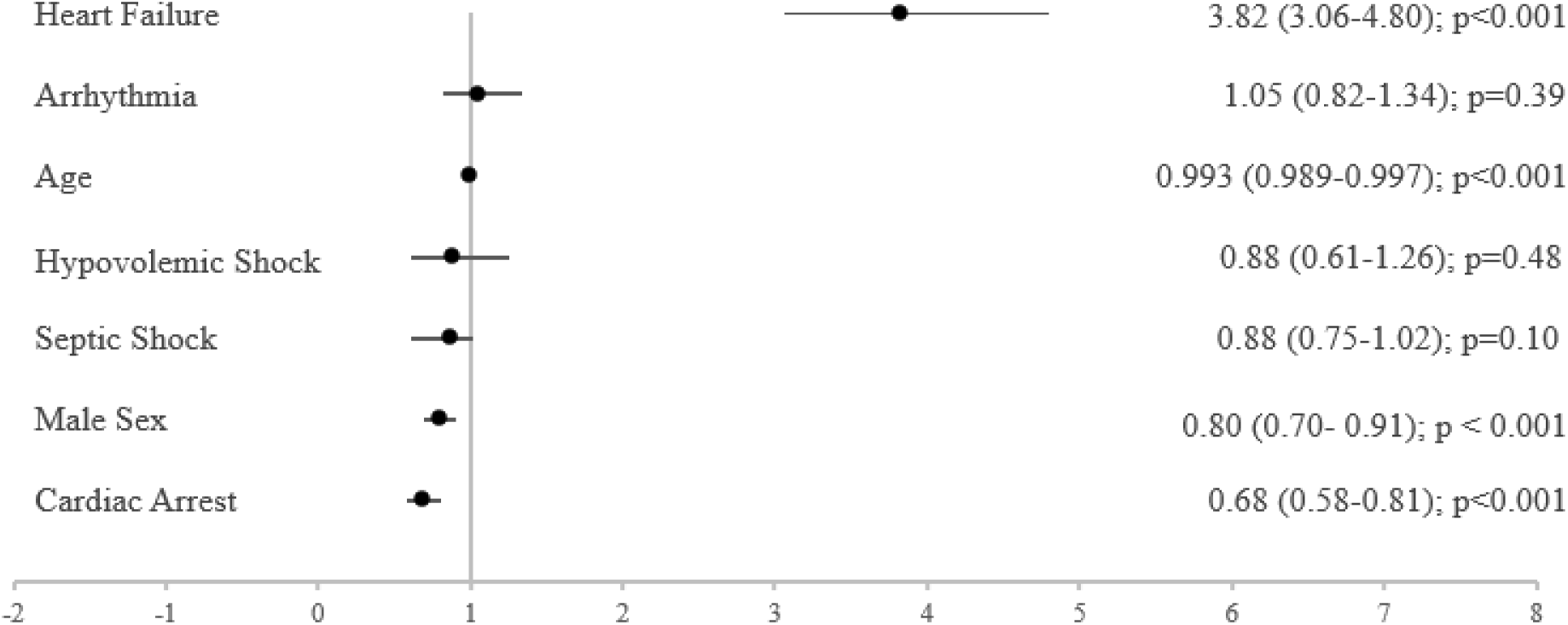
Odds ratios describing the likelihood of inodilator use across ICU subtype

The patients’ ICU subtype was found to be the strongest indicator of inodilator utilization in mixed effect modeling, as ICU location accounted for 15.8% of the variation. In comparison, patient-level characteristics combined accounted for a total of 6.1% of the variance in inodilator use. Of the identified patient-level variables, congestive heart failure accounted for the most significant proportion of variance, accounting for 4.0% of the 6.1% variance. 78.1% of the variance in inodilator use was not explained by the evaluated patient-level factors or by ICU location in this model.

## Discussion

In this retrospective analysis, we found (1) vasoactive medication utilization in cardiogenic shock is common and varied significantly amongst ICU environments within a single institution, (2) inodilator utilization in cardiogenic shock is common and varied significantly amongst ICU environments within a single institution, (3) inodilator utilization is impacted by patient-level factors, and (4) ICU environment impacts the frequency and choice to use an inodilator and accounts for more variation in usage than patient-level factors.

Our study cohort had high overall use of vasoactive medications with 91.5% of patients placed on at least one vasoactive medication during the hospital admission. This is a unique analysis of the frequency of vasoactive medication utilization for the treatment of cardiogenic shock, as it involves patients across all hospital settings, rather than isolating to a specific subtype of ICU, and only includes patients who have been diagnosed with cardiogenic shock. Norepinephrine was utilized most frequently in 77.1% of cases, which is consistent with prior trends suggesting norepinephrine has become the most frequently used vasoactive medication in the modern cardiac ICU.^21,22^ Other vasoactive medications were used less frequently but also exhibited broad variability in use across different ICU environments. The specific rationale for the selection of these medications was not evaluated and warrants further investigation in future research.

When considering identified variance in inodilator utilization captured by our model, the impact of patient factors, such as cardiac arrest and heart failure, likely represents tailored decision-making regarding inodilators to patient-level differences, which remain important to consider. In these situations, it is important to phenotype the subtype of cardiogenic shock to determine if use of inodilators would provide a meaningful benefit that outweighs any potential harms in the use of these medications. Despite the importance of this tailored decision-making, the patient-level factors identified in our model account for a smaller portion of the overall variance in usage of inodilators, raising the question whether these important considerations are driving the decisions surrounding the use of these medications, or if there are external, non-patient-level factors contributing significantly to the decision to use inodilators.

Specifically, in our model, the non-patient-level variable of ICU location explained more than 15% of the variance in the choice to use an inodilator. This individual factor accounted for more variance than all patient-level characteristics combined. These findings raise the important question of why ICU location accounts for such a significant effect on inodilator utilization. Ideally, the use of specific agents may represent customized care, but it may also represent clinical inertia and cognitive biases or provider comfort level in using these medications. In specialized cardiac and cardiothoracic ICUs, providers may be more accustomed to using inodilator medications, thus inodilators may be more likely to be used earlier in the treatment course, while in other ICUs where inodilators are less frequently used, and cardiogenic shock is less commonly managed, there may be more hesitation in initiating these treatment options. Beyond specific provider decision making, each ICU is likely to have different policies and procedures and training amongst its support staff for use of both pharmacologic interventions, like initiation of inodilators, as well as non-pharmacologic interventions, like mechanical circulatory support, which also may impact the availability of these treatment options. How this ultimately impacts patient outcomes and care was not evaluated in this study; however, it should be considered in further research. Addressing these and any other potential barriers to the implementation of therapeutic options may allow for more efficient and timely care for patients.

Additionally, the use of dobutamine and milrinone varied dramatically, with dobutamine preferred in the cardiac ICU, while milrinone was more commonly used in the cardiothoracic surgical ICU. This significant variation exists despite existing literature stating that neither dobutamine or milrinone was superior in management of cardiogenic shock.^5^ Further evaluation regarding factors within each ICU that drive choices of specific inodilator utilization is needed to completely understand this practice variation and guide further recommendations in the management of care.

Our study does have several limitations. First, it relies on large volume data extraction, and missing information or incorrectly used billing codes may introduce inaccuracy. In our study, if the medication or comorbidity was not retrieved from the clinical data repository, it was considered to be absent, and there is a possibility that the variable in question was mislabeled.

Secondly, our diverse patient sample spanned many years but was limited to a single center, and thus, it is unclear how generalizable the results are to other hospital systems or clinical environments. It is also possible that clinical practice patterns have evolved over the years, and the ten-year snapshot may not represent current practice patterns.

Finally, our mixed-effect model highlighted key factors associated with variance in vasoactive medication utilization for the treatment of cardiogenic shock, such as heart failure history and ICU location. Still, a considerable proportion of the variability in inodilator use (78.1%) remained unexplained. In selecting fixed effects, we aimed to minimize collinearity and reduce the risk of overfitting, thereby enhancing the model’s interpretability and generalizability. This unexplained variance suggests there may be unmeasured clinical or institutional factors that also contribute to variability.

## Conclusion

In this study, we demonstrated that the use of vasoactive medications, and more specifically inodilators, varies significantly across ICU subtypes within an institution. ICU location was found to account for a significant percentage of variance in inodilator utilization.

## Data Availability

All data produced in the present study are available upon reasonable request to the authors

## Acknowledgements

MM received funding support from the US National Institutes of Health (NHLBI, R01HL167790) paid to the University of Michigan, related to the present work. Additionally, MM has received a research grant from the US National Institutes of Health (NIDDK) and support from Chiesi, USA, paid to the University of Michigan, and unrelated to this present work. AD is supported by NIH-NHLBI [K08HL163328, 1R01HL167524]. AD is supported by the protein folding disease initiative, Michigan Biology of Cardiovascular Aging (M-BoCA) at the University of Michigan. AD receives compensation as editor for Merck Manuals.

## Disclosures

Dr. Mathis received funding support from the US National Institutes of Health (NHLBI, R01HL167790) paid to the University of Michigan, related to the present work. Additionally, Dr. Mathis has received a research grant from the US National Institutes of Health (NIDDK) and support from Chiesi, USA, paid to the University of Michigan, and unrelated to this present work. Dr. Thompson is supported by NIH-NHLBI [K08HL163328, 1R01HL167524]. Dr. Thompson is supported by the protein folding disease initiative, Michigan Biology of Cardiovascular Aging (M-BoCA) at the University of Michigan. Dr. Thompson receives compensation as editor for Merck Manuals.

## Notes

### Funding Statement

This study did not receive any funding

